# The impact of SARS-CoV-2 transmission fear and COVID-19 pandemic on the mental health of patients with primary immunodeficiency disorders, severe asthma, and other high-risk groups

**DOI:** 10.1101/2020.06.26.20140616

**Authors:** Fatih Çölkesen, Oğuzhan Kılınçel, Mehmet Sözen, Eray Yıldız, Şengül Beyaz, Fatma Çölkesen, Gökhan Aytekin, Mehmet Zahid Koçak, Yakup Alsancak, Murat Araz, Şevket Arslan

**Author notes:** **Corresponding Author:** Fatih Çölkesen, Division of Clinical Immunology and Allergy, Department of Internal Medicine, Meram Faculty of Medicine, Necmettin Erbakan University, Abdulhamid Han Avenue, 3 42090, Meram, Konya, Turkey.

## Abstract

**Background:** The adverse effects of COVID-19 pandemic on the mental health of high-risk group patients for morbidity and mortality and its impact on public health in the long term have not been clearly determined.

**Objective:** To determine the level of COVID-19 related transmission fear and anxiety in healthcare workers and patients with primary immunodeficiency disorder (PID), severe asthma, and the ones with other comorbidities.

**Methods:** The healthcare workers and patients with PID, severe asthma (all patients receiving biological agent treatment), malignancy, cardiovascular disease, hypertension (90% of patients receiving ACEI or ARB therapy), diabetes mellitus (42 % of patients receiving DPP-4 inhibitor therapy) were included in the study. A total of 560 participants, 80 individuals in each group, were provided. The hospital anxiety and depression scale (HADS) and Fear of illness and virus evaluation (FIVE) scales were applied to the groups with face to face interview methods.

**Results:** The mean age was 49.30 ± 13.74 years and 306 (55 %) were female. The FIVE Scale and HADS-A scale scores of health care workers were significantly higher than other groups scores (p = 0.001 and 0.006). The second-highest scores belonged to patients with PID. There was no significant difference between the groups for the HADS-D score (p=0.07). The lowest score in all scales was observed in patients with hypertension.

**Conclusions:** This study demonstrated that in the pandemic process, patients with primary immunodeficiency, asthma patients, and other comorbid patients, especially healthcare workers, should be referred to the centers for the detection and treatment of mental health conditions.

## Introduction

Severe acute respiratory syndrome coronavirus 2 (SARS-CoV-2) and the disease it causes, coronavirus disease 2019 (COVID-19), became pandemic worldwide in 2020. When we started to write the article (May 25, 2020), the total number of cases in the affected 215 countries of the world was 5.304.772, and the total number of deaths was 342.029. The total number of cases was 156.827, and the total number of deaths in Turkey in 4340 (1, 2). The spectrum of symptomatic infection ranges from mild to fatal. Pneumonia is the most common serious infection, characterized by fever, dry cough, dyspnea and bilateral infiltrates in chest imaging (3, 4). COVID-19 also occurs in adults with severe disease in predominantly older patients or patients with underlying medical comorbidities. Comorbidities associated with severe disease and mortality include; primary or secondary immunodeficiency disease, pre-existing pulmonary disease, cardiovascular disease, diabetes mellitus, hypertension, and malignancies (3, 5-7).

The main way of transmission and spread of the infection is with the respiratory tract and in the form of clumps especially with close contact near the environment. Due to the acute nature of the pandemic and the spread and infectious power of the virus, it will undoubtedly cause anxiety, depression, and other psychological disorders in humans (8). People without immune system defects can recover even with Covid-19 disease, but this may not be possible in patients with primary and secondary immunodeficiency. Patients with primary immunodeficiency, whose health-related quality of life is lower than healthy people, will particularly need mental health support in this process (9). Taking into account the psychological status and mental health support of primary immunodeficiency patients, severe asthma patients, and other patients with comorbidities (malignancy, cardiovascular system diseases, diabetes mellitus, hypertension), which are the high risk patient groups in terms of morbidity and mortality in the COVID-19 pandemic process, is crucial.

In our study, we compare COVID-19 transmission fear induced anxiety and depression in patients with primary immunodeficiency disorders who are naturally susceptible to infections, between other comorbid patients and those healthcare workers in the frontline of the COVID-19 pandemic.

## Methods

### Objective, Study design and Setting

#### Objective of the study

To determine the level of COVID-19 related transmission fear and anxiety in PID patients, severe allergic asthma and severe eosinophilic asthma patients, patients with other comorbidities,and health workers. Thus, to reveal the necessity of supportive psychological treatments, which are ignored simultaneously by giving full attention to improving vaccines and other therapies that control infection during the pandemic process.

#### Study design

After the first case of COVID-19 was diagnosed on March 11 2020, a pandemic action plan was initiated in all units at Necmettin Erbakan University Meram Faculty of Medicine. Hospital buildings are divided into two parts: pandemic hospital and non-pandemic patient care hospital. Treatment of patients with primary immunodeficiency and other patients continued at the non-pandemic hospital. The common view of immunoglobulin therapy in our clinical immunology clinic is SCIG therapy in patients who can adapt since the majority of our patients with primary immunodeficiency come from remote areas. Before the pandemic, SCIG treatment hands-on training was given to all patients who were eligible for the clinic, but only half were able to adapt. Total 80 primary immunodeficiency patients (58 CVID, 5 CID, 2 Wiskott Aldrich syndrome, 3 Hyper Ig E syndrome, 8 symptomatic isolated NK deficiency, 3 Chronic Granulomatous Disease, 1 Bloom Syndrome), 80 patients with severe asthma (63 patients receiving Omalizumab therapy, 17 patients receiving Mepolizumab therapy), 80 HT patients (72 patients receiving ACEI or ARB therapy), 80 DM patients (38 receiving DPP-4 inhibitor therapy), 80 patients with cardiovascular system diseases (42 Arrhythmia Patients, 26 congestive heart failure, 12 patients with coronary artery disease), 80 malignancy patients (22 colorectal carcinomas, 18 breast carcinomas, 17 lung carcinomas, 5 prostate carcinomas, 4 pancreatic carcinomas, 4 endometrium carcinomas, 3 gastric carcinomas, 3 cervix uteri carcinomas, 3 bladder carcinomas, 1 HCC) and 80 health-care workers (36 doctors, 20 nurses, 13 patient caregivers, 6 cleaners, 5 medical secretaries) actively working to combat the COVID-19 pandemic were included in the study. We started the study on April 20, and we closed the survey on May 15. Verbal informed consent was obtained from all participants. Hospital Anxiety and Depression Scale (HADS) and Fear of Illness and Virus Evaluation (FIVE) scales were applied to the groups with face to face interview methods. The results of the groups were compared with each other.

#### Setting

Questionnaires of patients with primary immunodeficiency and severe asthma patients who are receiving mepolizumab or omalizumab therapy were performed at the Clinical Immunology and Allergy Department of Meram Faculty of Medicine, Necmettin Erbakan University, Konya/Turkey. Questionnaires of patients with malignancy were performed in the Clinical Oncology Department, the questionnaires of patients with cardiovascular system disease and hypertension patients were organized in the Cardiology department of the same faculty. The questionnaires of diabetes patients were conducted in the Department of Endocrinology and metabolism diseases of Kocaeli University Faculty of Medicine. In these two hospitals, questionnaires were applied to healthcare workers who are actively working to combat the COVID-19 pandemic. The prevalence of COVID-19 infection was high in both hospitals.

### Questionnaires

#### Fear of Illness and Virus Evaluation (FIVE) Scale

The Fear of Illness and Virus Evaluation (FIVE) Scale was created by Prof. Dr. Jill Ehrenreich-May from Miami University. The scale translated to Turkish and firstly used by Dr. Zekiye Çelikbaş from Gaziosmanpaşa University. The scale has 3 forms: Adult, Child, and Parent form. Adult form was used in our study. The answers are scored in a 4-point Likert format and 1-4. The scale consists of 4 parts: Fears about Contamination and Illness (9-item, 9-36 score ranging), Fears about Social Distancing (10-item, 10-40 score ranging), Behaviors Related to Illness and Virus Fears (14-item, 14-56 score ranging), Impact of Illness and Virus Fears (2-item, 2-8 score ranging). There are a total of 35 items on the scale and the total score ranges from 35 to 140. In the use of the scale, permission was obtained from those who prepared both the original and Turkish forms.

#### Hospital Anxiety and Depression Scale

The scale was developed by Zigmond and Snaith (10). It is used to screen depression and anxiety in those with medical illnesses. The scale consists of 14 items; 7 of them assess anxiety and 7 of them assess depression. Answers are scored between 0-3 in quadruple Likert formats. The lowest score that patients can get from both subscales (anxiety and depression subscale) is 0 and the highest score is 21. Turkish reliability and validity were done by Aydemir et al. The Turkish version of the HAD scale has been found to be valid and reliable in medical patients (Cronbach’s α of 0.8525 and 0.7784 for the HAD anxiety subscale and depression subscale respectively). In the Turkish version of the HAD scale, the cut-off score for anxiety subscale was found to be 10 and 7 for depression subscale (11).

### Statistical analysis

SPSS version 22.0 statistical package software (IBM Corp., Armonk, NY, United States) was used for statistical analyses. Continuous variables are demonstrated as mean ± standard deviation, median (min-max), and categorical variables as numbers and percentages. Kolmogorov–Smirnov test was used for evaluating the normality of distribution. When parametric test assumptions are provided, Independent-Samples T-Test and One-way ANOVA test, when parametric test assumptions are not provided, Mann-Whitney U test and Kruskal-Wallis test were used to compare independent group differences. The linear relation between the continuous variables was evaluated using Pearson (r) correlation analysis. ROC analysis method was used for diagnostic performance analysis of variables. The chi-square test was performed to compare the study groups in terms of categorical variables. The threshold for significance was defined at p < 0.05.

## Results

Patients with PID, severe asthma, malignancy, CVS disease, HT, DM, and healthcare workers were included in the study. It provided 560 participants, including 80 from each group. In this study, there were 306 (55%) female and 254 (45%) male subjects. When the groups were evaluated separately, there was no statistically significant difference in terms of gender and age (p = 0.08 and 0.46) (Table 1).

**Table 1.**
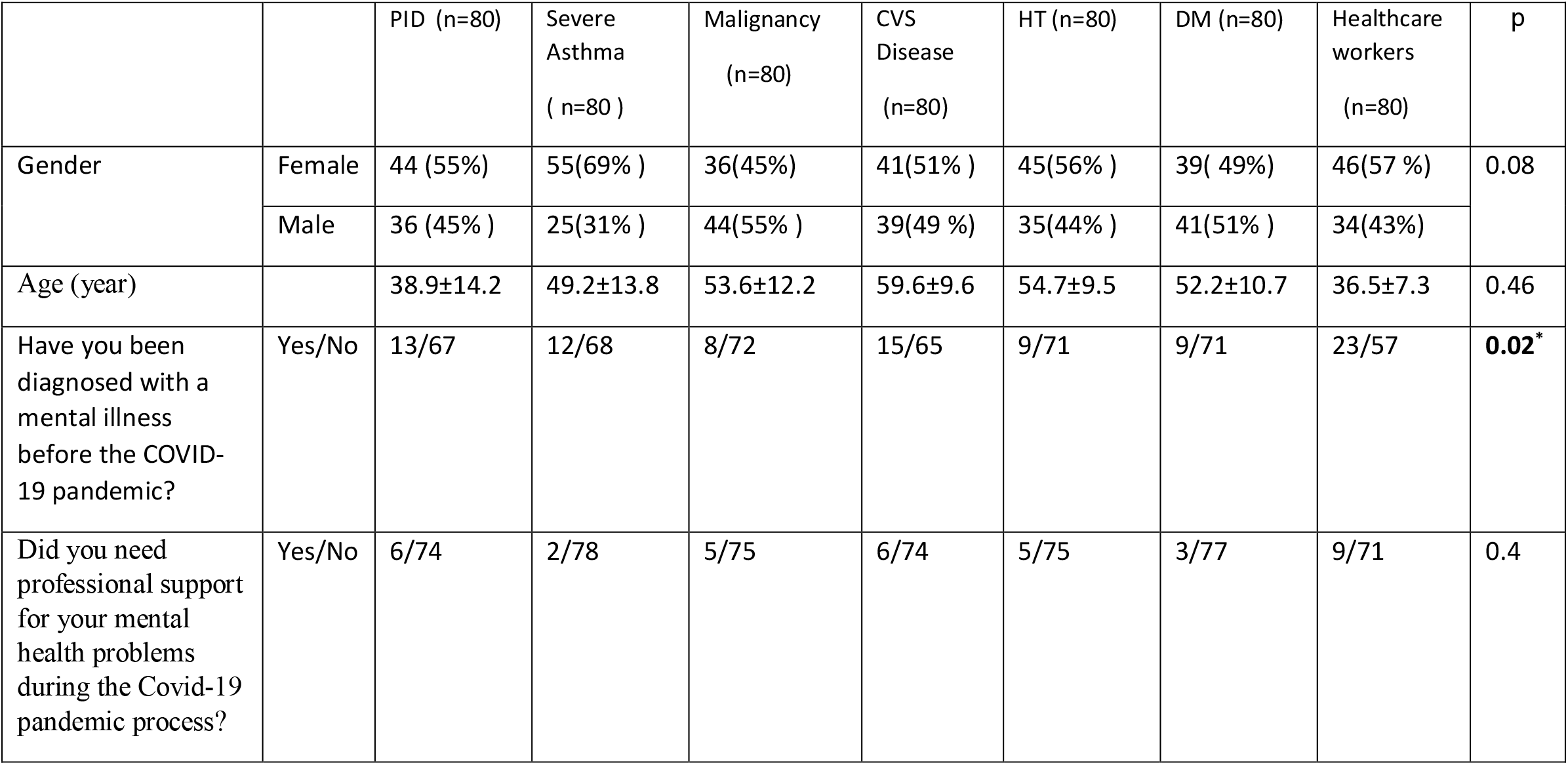
Age, Gender and Mental Health Stories of the Participants

|  |  | PID (n=80) | Severe Asthma<br>( n=80 ) | Malignancy<br>(n=80) | CVS Disease<br>(n=80) | HT (n=80) | DM (n=80) | Healthcare workers<br>(n=80) | p |
| --- | --- | --- | --- | --- | --- | --- | --- | --- | --- |
| Gender | Female | 44 (55%) | 55(69% ) | 36(45%) | 41(51% ) | 45(56% ) | 39( 49%) | 46(57 %) | 0.08 |
|  | Male | 36 (45% ) | 25(31% ) | 44(55% ) | 39(49 %) | 35(44% ) | 41(51% ) | 34(43%) |  |
| Age (year) |  | 38.9±14.2 | 49.2±13.8 | 53.6±12.2 | 59.6±9.6 | 54.7±9.5 | 52.2±10.7 | 36.5±7.3 | 0.46 |
| Have you been diagnosed with a mental illness before the COVID-19 pandemic? | Yes/No | 13/67 | 12/68 | 8/72 | 15/65 | 9/71 | 9/71 | 23/57 | **0.02\*** |
| Did you need professional support for your mental health problems during the Covid-19 pandemic process? | Yes/No | 6/74 | 2/78 | 5/75 | 6/74 | 5/75 | 3/77 | 9/71 | 0.4 |

**Table 2.**
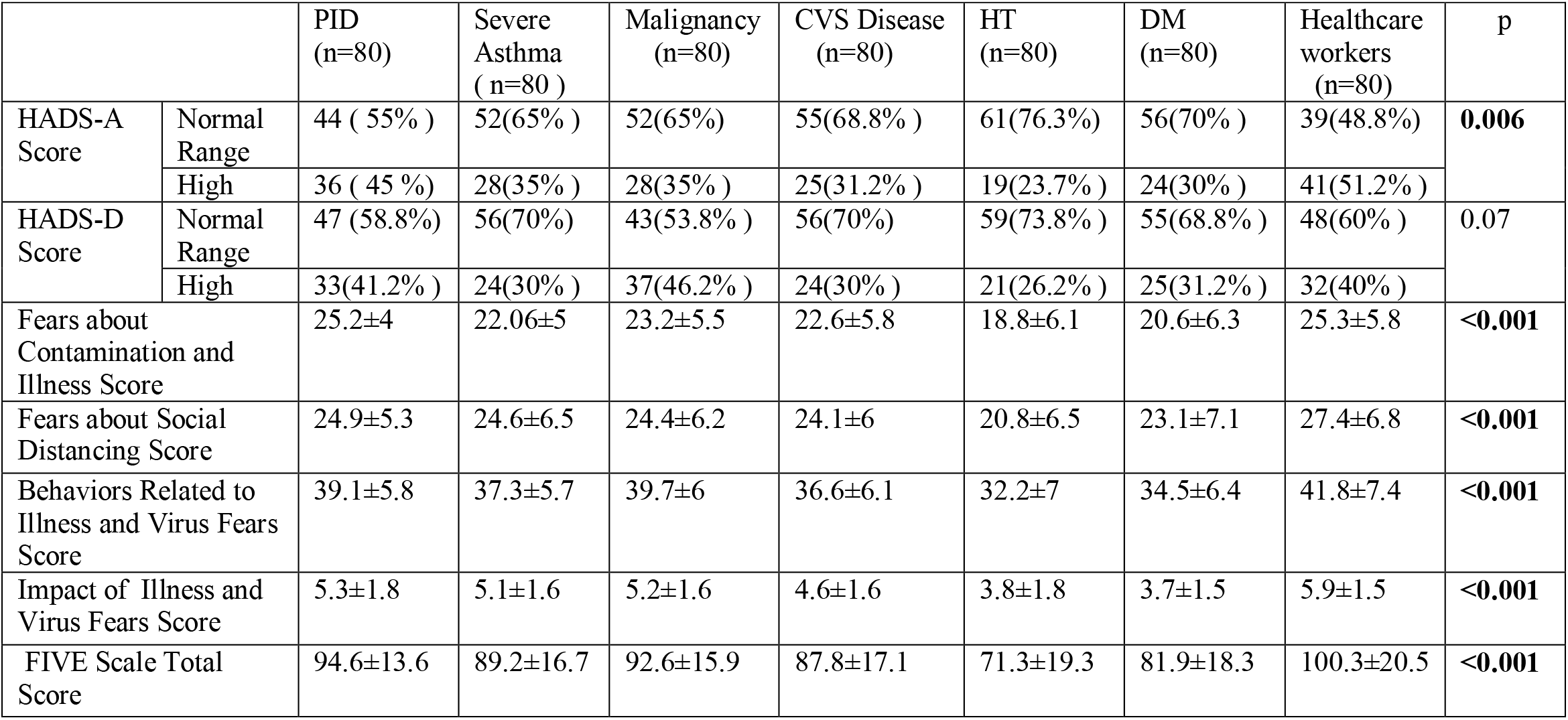
Participant Groups’ Hospital Anxiety and Depression Scale (HADS) Scores, Fear of Illness and Virus Evaluation (FIVE) Scale Scores

In the period of before the COVID-19 pandemic, the number of subjects who received professional support for their mental problem was the highest in the healthcare workers group (28.75%, 23/80). In other groups, this rate was as follows: 16.25% (13/80) in PID group, 15% (12/80) in severe asthma group, 10% (8/80) in malignancy group, 18.75% in CVS disease group (15 / 80), 11.25% (9/80) in the HT group and 11.25% (9/80) in the DM group (p = 0.02). Considering those who received professional support for their mental problems during the COVID-19 pandemic process, the highest rate was again determined in the healthcare workers group (11.25 %, 9/80, p = 0.4) (Table 1).

Regarding the evaluation of all participants, there were a strong positive correlation between FIVE scale scores and anxiety (r=0.828; p<0.001) (Figure 1) and pearson correlation analysis showed a moderate positive correlation between FIVE scale scores and depression (r = 0.660; p<0.001) (Figure 2). The effectiveness of FIVE scales in distinguishing participants with and without anxiety; the scale’s cut-off total score was 96, with 79.1 % sensitivity and 86.6 % specificity. FIVE scale was found to have a significant discrimination power. (AUC = 0.870, p <0.0001, 95% CI (lower bound – upper bound) = 0.836 - 0.904) (Table 3). The ability of the FIVE scales to distinguish participants with and without depression was significant, though not as high as in anxiety. When scale cut-off total score 96 was taken, sensitivity was 62.8 % and specificity was 76.9 % (AUC = 0.760, p <0.0001, 95% CI (lower bound – upper bound) = 0.717 - 0.803) (Table 4).

**Table 3.** Determination of the ability of Fear of Illness and Virus Evaluation (FIVE) scale scores to predict COVID-19 related anxiety through ROC curv

| Variables | AUC ( % 95 CI ) | Cut-off | p | Sensitivity (%) | Specificity (%) |
| --- | --- | --- | --- | --- | --- |
| FIVE PART-1 SCORE | 0,833 (0,797 - 0,868) | 24 | < 0.001 | 78.6 | 76.6 |
| FIVE PART-2 SCORE | 0,843 (0,808 - 0,878) | 26 | < 0.001 | 73.6 | 78.6 |
| FIVE PART-3 SCORE | 0,795 (0,756 - 0,835) | 40 | < 0.001 | 71.1 | 79.4 |
| FIVE PART-4 SCORE | 0,857 (0,824 - 0,889) | 6 | < 0.001 | 77.1 | 83,3 |
| FIVE TOTAL SCORE | 0,870 (0,836 - 0,904) | 96 | < 0.001 | 79.1 | 86.6 |

**Table 4.** Determination of the ability of Fear of Illness and Virus Evaluation (FIVE) scale scores to predict COVID-19 related depression through ROC curve

| Variables | AUC ( % 95 CI ) | Cut-off | p | Sensitivity (%) | Specifity (%) |
| --- | --- | --- | --- | --- | --- |
| FIVE PART-1 SCORE | 0,737 (0,693 - 0,781) | 24 | < 0.001 | 67.3 | 69.8 |
| FIVE PART-2 SCORE | 0,748 (0,704 - 0,791) | 26 | < 0.001 | 64.8 | 73.1 |
| FIVE PART-3 SCORE | 0,705 (0,658 - 0,751) | 40 | < 0.001 | 60.7 | 73.1 |
| FIVE PART-4 SCORE | 0,780 (0,739 - 0,821) | 6 | < 0.001 | 68.9 | 78,0 |
| FIVE TOTAL SCORE | 0,760 (0,717 - 0,803) | 96 | < 0.001 | 62.8 | 76.9 |

**Figure 1.**
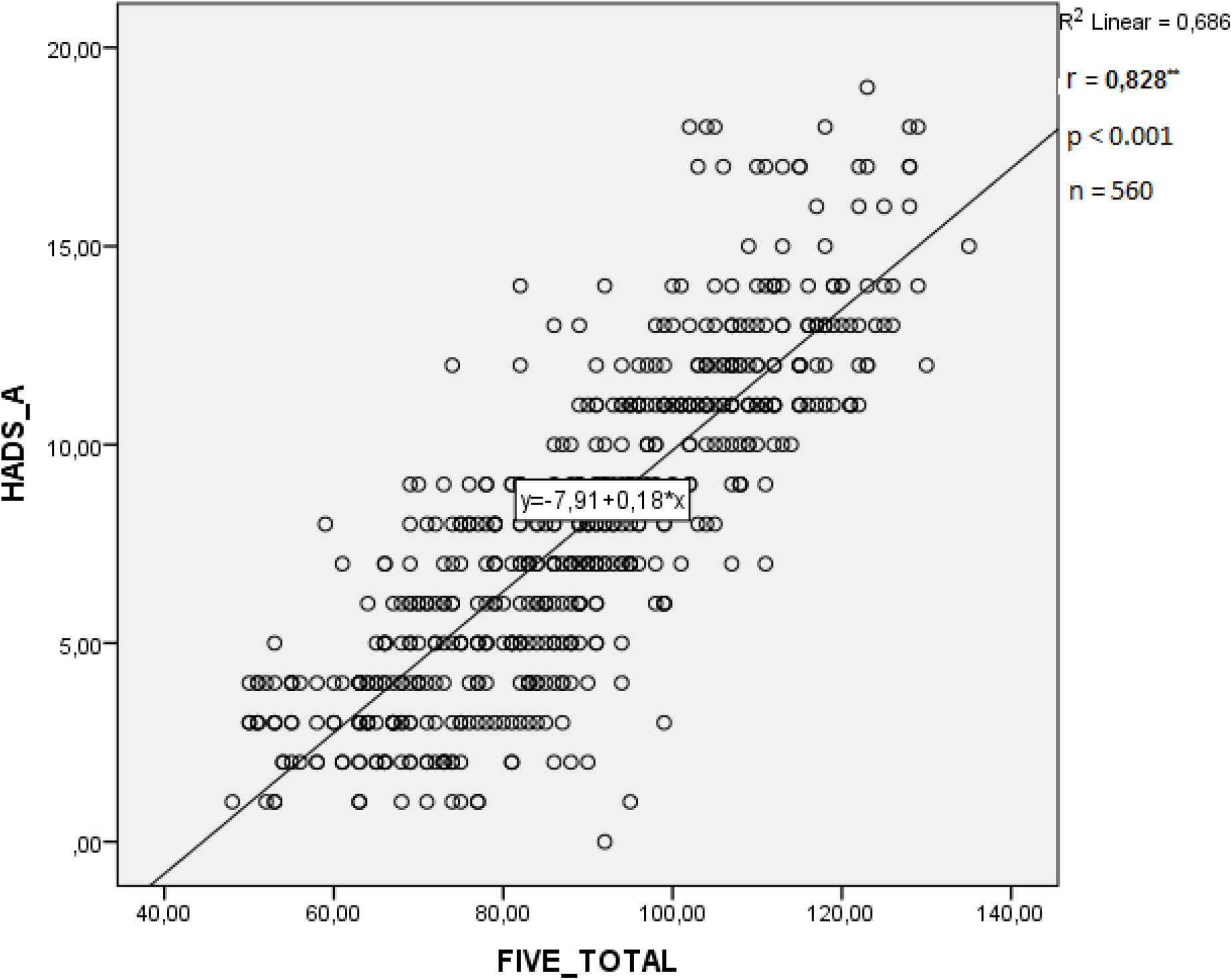
Pearson correlation analysis showed a strong positive correlation of FIVE_TOTAL vs HADS_A (Pearson r = 0.828; p < 0.001; n = 560). Line represents linear regression of data (y = -7.91+ 0.18*x; r^2^ = 0.686). Abbreviations: FIVE_TOTAL, Fear of Illness and Virus Evaluation Scale Total Score; HADS_A, Hospital Anxiety and Depression Scale, Anxiety Subscale Score.

**Figure 2.**
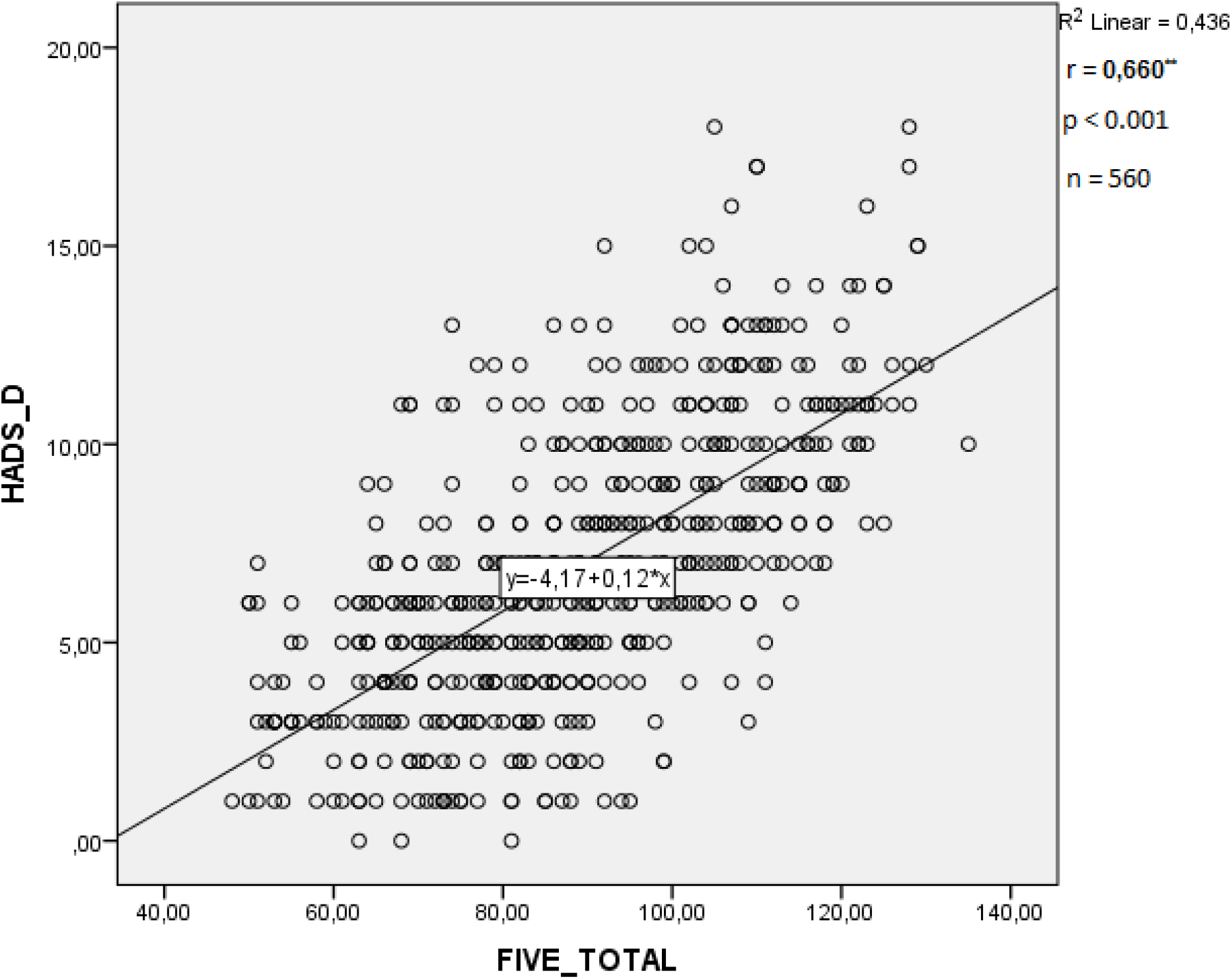
Pearson correlation analysis showed a moderate positive correlation of FIVE_TOTAL vs HADS_D (Pearson r = 0.660; p < 0.001; n = 560). Line represents linear regression of data (y = -4.17 + 0.12*x; r^2^ = 0.436). Abbreviations: FIVE_TOTAL, Fear of Illness and Virus Evaluation Scale Total Score; HADS_D, Hospital Anxiety and Depression Scale, Depression Subscale Score.

**Figure 3.**
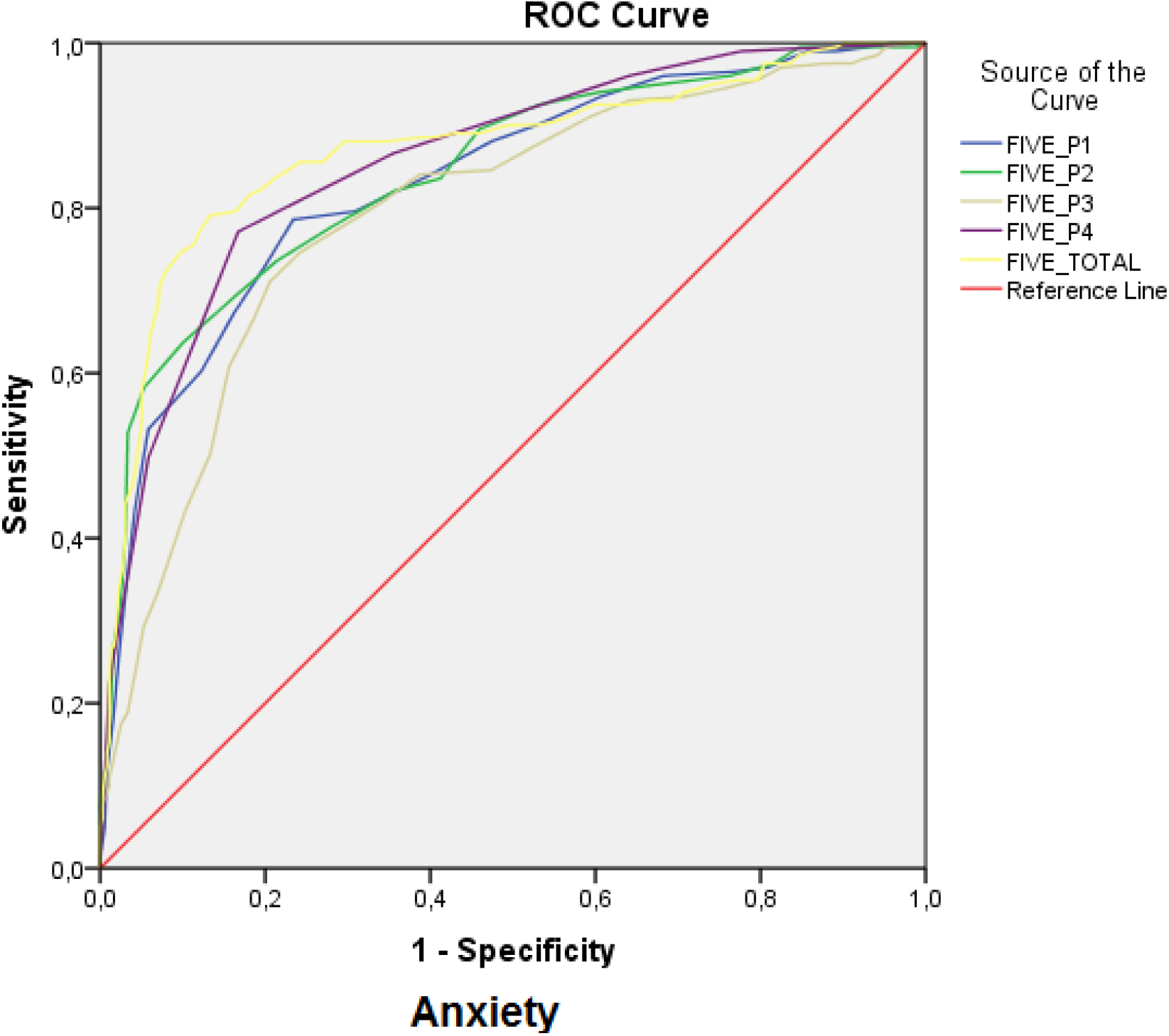
ROC analysis of FIVE Scale Total Score and parts of scale scores (FIVE P1 to P4) baseline values for anxiety. Notes: FIVE Scale Total Score and parts of scale scores were set to a positive influence, and specificity and sensitivity of FIVE Scale Total Score and parts of scale scores were plotted. **Abbreviations:** ROC, receiver operating characteristic; FIVE, Fear of Illness and Virus Evaluation; FIVE P1, FIVE Part-1(Fears about Contamination and Illness); FIVE P2, FIVE Part-2 (Fears about Social Distancing); FIVE P3, FIVE Part-3 (Behaviors Related to Illness and Virus Fears); FIVE P4, FIVE Part-4 (Impact of Illness and Virus Fears), FIVE TOTAL, Fear of Illness and Virus Evaluation Scale Total Score.

**Figure 4.**
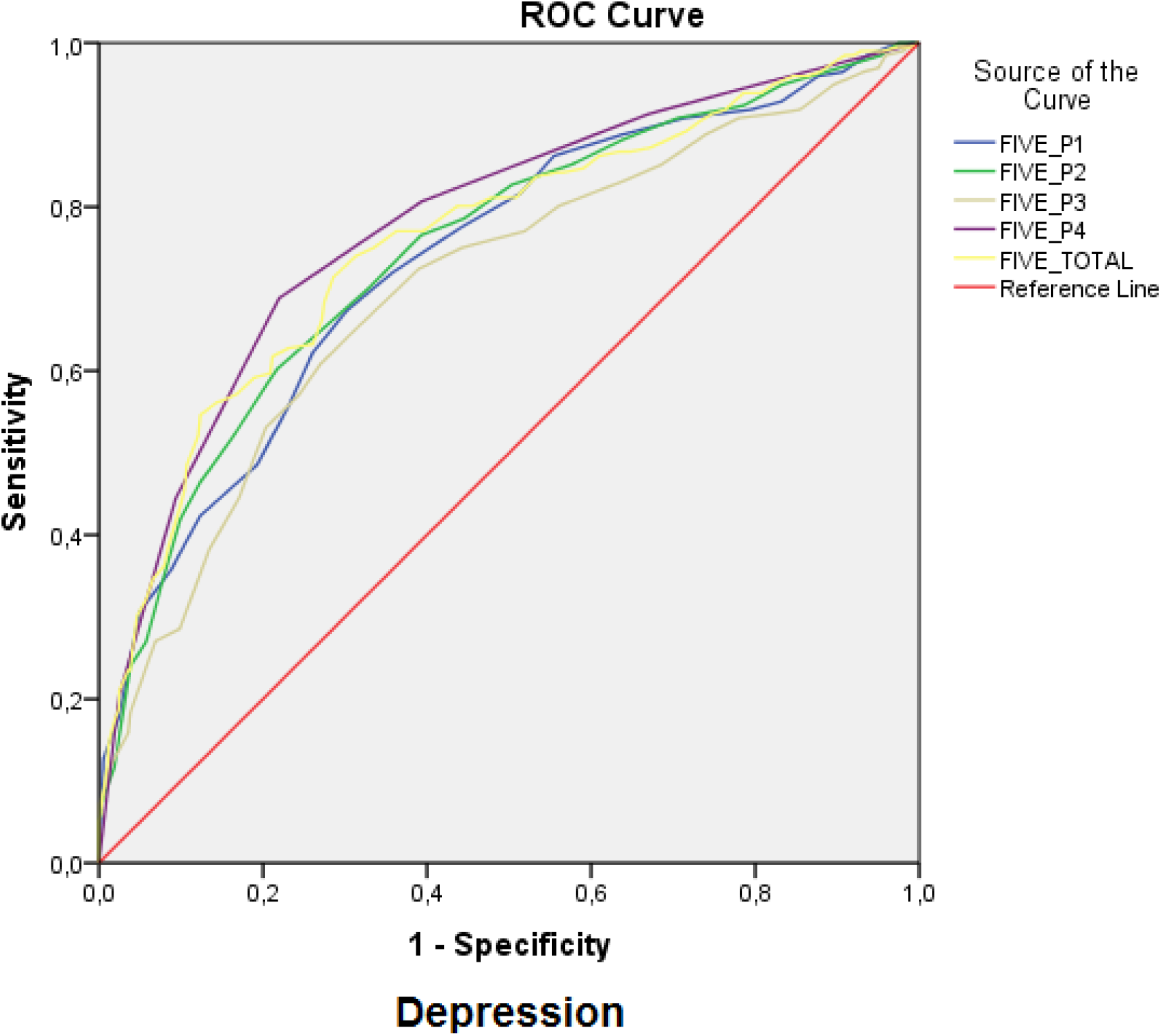
ROC analysis of FIVE Scale Total Score and parts of scale scores (FIVE P1 to P4) baseline values for depression. Notes: FIVE Scale Total Score and parts of scale scores were set to a positive influence, and specificity and sensitivity of FIVE Scale Total Score and parts of scale scores were plotted. **Abbreviations:** ROC, receiver operating characteristic; FIVE, Fear of Illness and Virus Evaluation; FIVE P1, FIVE Part-1(Fears about Contamination and Illness); FIVE P2, FIVE Part-2 (Fears about Social Distancing); FIVE P3, FIVE Part-3 (Behaviors Related to Illness and Virus Fears); FIVE P4, FIVE Part-4 (Impact of Illness and Virus Fears), FIVE TOTAL, Fear of Illness and Virus Evaluation Scale Total Score.

The healthcare workers group had the highest score in 4 subscales (Fears about Contamination and Illness, Fears about Social Distancing, Behaviors Related to Illness and Virus Fears, Impact of Illness and Virus Fears) and on the whole scale of FIVE (p <0.001 for all). The second group with the highest score after the health care workers (100.3±20.5) in the total scale score was the PID group (94.6 ± 13.6). The lowest score in the scale was observed in patients with hypertension (71.3 ± 19.3) (Table 2).

The participants were evaluated in terms of anxiety and depression according to the cut-off values (10 points for HADS-A and 7 points for HADS-D) determined in the Turkish validation study of the HADS scale. The group with the highest proportion of participants with a HADS-A score higher than the limit value was the healthcare workers (51.2%, 41/80) then respectively PID (45%, 36/80), malignancy (35%, 28/80) and severe asthma (35%, 28/80) group participants. The group with the lowest rate was the HT group (23.7%, 19/80), (p = 0.006) (Table 2).

In the HADS-D subscale, there was no statistically significant difference between groups (p=0.07). Nevertheless, the group with the highest score was the participants with malignancy (46.2%, 37/80). PID patients (41.2%, 33/80) and healthcare workers (40%, 32/80) were the other groups with the highest HADS-D subscale score. In this subscale, the lowest rate belonged to the HT group participants (% 26.8, 21/80) (p=0.07) (Table 2).

## Discussion

Survey results reported by patients in clinical practice have been proposed as a means of improving doctor-patient communication, revealing patients’ problems, screening functional problems (9). In the current study we applied a new scale, Fear of Illness and Virus Evaluation (FIVE). The FIVE scale was evaluated as a useful assessment method for the detection of anxiety and depression due to fear of disease and virus transmission. The results indicate that the FIVE scale and HADS-A scale scores of health care workers running to fight COVID-19 pandemic were significantly higher than those of the primary immunodeficiency patients and other comorbidity patient groups scores (p = 0.001 and 0.006). The second-highest score after healthcare workers belonged to patients with primary immunodeficiency. There was no significant difference between the groups in terms of the HADS-D score (p=0.07); on the other hand, the highest score belonged to patients with malignancy. The lowest score in all scales was observed in patients with hypertension.

Primary immune deficiency disorders are a group of more than 400 congenital immune defects that continue to expand with discovered novel defects (12). Some defects affect basic immunological pathways and result in susceptibility to both common and opportunistic pathogens, resulting in recurrent or chronic infections in most patients (13). In a study of children with primary immunodeficiency, a higher mental health disorder was detected in these patients than in children with chronic diseases such as severe asthma and chronic renal disease (14). These mental health disorders include depression, anxiety, somatization, social withdrawal and decreased social skills. Besides, 18% of pediatric-onset CVID patients had depression and were associated with mortality, especially in patients with delayed diagnosis (15). Patients with primary immunodeficiencies are more vulnerable to SARS-CoV-2, and the disease it causes COVID-19, similar to other infectious agents, compared to immune-competent individuals (9). So, it is inevitable that the COVID-19 pandemic impacts health-related quality of life (HRQoL) and the risk of anxiety/depression in patients with primary immunodeficiency. In patients with primary immunodeficiency, anxiety and depression were significantly higher compared to the healthy population. Mental disorders contribute to PID morbidity and mortality (15). In order to improve the quality of life in these patients, referrals should be made as soon as possible, and treatment should be started (16).

Severe asthma is defined by the presence of ≥ 3 of the following criteria: having >2 asthma attacks per week, having asthma-induced night awakenings, the constant need a reliever (short-acting beta-2 agonist) for controlling asthma symptoms, and extremely limited normal activation (17). Recent studies have shown that stress might increase the risk of asthma and asthma-related morbidity by affecting the immune system (18). Although asthma does not seem to be a severe risk factor for COVID-19, poorly controlled asthma can lead to a more complicated course of disease for patients with COVID-19 (19). However, in a recently published study authors reported that the most common comorbidities among young patients hospitalized for COVID-19 are asthma, diabetes, and obesity (20). Due to the role of asthma in COVID-19 prognosis uncertain yet, anxiety remains high in patients. Asthma is a susceptible disease to viral infections, and about 80% of asthma exacerbations are associated with viral infections. In allergic asthmatic patients, allergic sensitization and eosinophilic inflammation can disrupt the integrity of the airway epithelium. Thereby paving the way for limiting the ability of viruses clearance and foster the location of viruses in the lower respiratory tract. Therefore, it is thought that biologic agents, such as Omalizumab (an anti-IgE antibody) and Mepolizumab (a monoclonal antibody to IL-5), which are used in the treatment of severe allergic or eosinophilic asthma, may have positive effects on the prognosis of COVID-19 (21). Thus, the risk of COVID-19 related anxiety is also expected to be lower as virus induced asthma exacerbations will decrease as a result of using biological agents in these patients. The results of our study also support this theory.

There are some pandemic specific problems on the basis of higher FIVE scale scores and HADS-A scores in healthcare workers. One of the main causes of this distress in healthcare workers is the fear of being infected with the virus and spreading it to their families (22). This fear requires isolation from their families and they are also deprived of family support. Changes in the workplace, increase in working hours and workload are other factors that negatively affect the mental health of healthcare workers. In addition, social stigmatization and exclusion behaviors towards healthcare workers, who are considered to be the most exposed to the virus by the public, contribute to mental stress (23). The increase in the number of cases and mortality rates, as well as witnessing critical illnesses and deaths of their colleagues increase the mental breakdown (24). The shortage of personal protective equipment (PPE) and other materials is one of the essential reasons that increase anxiety for transmission (25). It is essential to take urgent measures to protect the mental health of healthcare workers and the smooth running of health services. The measures that can be taken in this regard are:

- Working hours should be arranged; breaks should be planned by considering physical and mental health.
- Personal protective equipment should be supplied in sufficient numbers, and a sense of trust should be created in the employees.
- Frontline employees should be changed at certain intervals to share risk
- Family, friends should be allowed to support (education of relatives of health professionals should be provided)
- Rewards should be made
- The detected mood changes should be treated at an early stage, without turning into permanent psychological disorders.

In a recent study from Wuhan, China, severe symptoms, need for mechanical ventilation, and risk of death was higher in patients with malignancy compared to COVID-19 patients without cancer (26). Psychiatric disorders such as major depression are more common in patients with malignancy compared to the general population (27). Depression is often accompanied by anxiety in these patients (28). It is crucial to support patients with malignancy and improve the quality of life, who have a predisposition to mental health disorders, against the adverse psychological effects of the COVID-19 pandemic process (29).

Patients with diabetes mellitus, cardiovascular system disease, and hypertension have been demonstrated to be associated with an increased risk of severe disease and mortality risk for COVID-19 (30-32). SARS-CoV-2 enters the cell by binding to the ACE2 surface receptor. The SARS-CoV-2 spike protein binds directly to the cell surface ACE2 receptor of the host cell, thereby facilitating the entry and replication of the virus into the cell (33). Based on this information, speculation about renin-angiotensin system inhibitors that may increase ACE2 levels and the use of these drugs will adversely affect the prognosis of COVID-19, which has ended with multicentre and extensive patient studies. The same applies to speculation between DPP-4 inhibitor drugs and diabetes mellitus prognosis concerning the COVID-19 (34). In our study, where we evaluated the mental health of comorbidity patients with high risk in terms of COVID-19 prognosis, the group with the lowest scores were patients with hypertension. However, according to our findings, even in patients with hypertension who received the lowest scale scores and relatively better than other patient groups, only a quarter of patients who need psychological support are still receiving treatment.

Nowadays, the focus is on drugs and vaccine discovery for the eradication of COVID-19, ignoring the mental health status of healthcare professionals, patients with primary immunodeficiency, asthma patients and other comorbidity patient groups, will have important implications for the community in the long run. Authorities and clinicians should provide support and take precautions in this regard before time.

The present study has several limitations. First, only a part of the participants had official diagnoses obtained by examining mental health professionals. Patients who were found meaningful in terms of mental health disorders with scales evaluation were referred to the psychiatry clinic. However, due to the harmful effects of the pandemic process, information feedbacks were not received. Second, since the study was performed during the pandemic process and in a hospital setting by face-to-face interview method, it was not compared with the mental health of the control group without comorbidity from the general population. Finally, it cannot be denied that the face-to-face interview method between the participants and the physicians in hospital settings may impact individuals’ mental health in the pandemic process. Even if the COVID-19 patients were not followed in these clinics and special measures were taken for the care of other patients.

In our knowledge, the current study is the first article to compare fear of infection transmission related anxiety and depression in adults with primary immunodeficiency and other high-risk group patients. Also, no published study has been found in the literature on the similar subject of the COVID-19 pandemic. We believe that the study will increase knowledge, especially in determining the anxiety and depression levels of patients with primary immunodeficiency, to be treated and to improve their quality of life.

## Conclusion

This study demonstrated that in the pandemic process, patients with primary immunodeficiency, asthma patients, and other comorbid patients, especially healthcare workers, should be referred to the centers for the detection and treatment of mental health conditions. Due to the mental disorders caused by the COVID-19 pandemic, the authorities should take precautions to prevent healthcare services from being interrupted and prevent harmful effects on the general population’s mental health.

## Data Availability

All data referred to in the manuscript is available.

## Acknowledgements

We would like to thank Prof. Dr. Jill Ehrenreich-May from Miami University for creating the FIVE scales, and Dr. Zekiye Çelikbaş from Gaziosmanpaşa University for the Turkish version of the scale. We thank all patients for their participation in the study. We would like to thank all healthcare professionals working in the COVID-19 pandemic process and all scientists who have contributed to the diagnosis, treatment, and management of the pandemic through clinical trials.

## Ethical Approval

The study was approved by the local ethics committee of Necmettin Erbakan University, Meram Faculty of Medicine with the 2020/2448 ID number, and the study was conducted according to the 1975 Declaration of Helsinki.

## Informed Consent

Informed consent was obtained from all participants.

## Author Contributions

All authors contributed to the design of the study. F.Ç^1^. and O.K. determined the scales used in the study. ş.A. applied scales to primary immunodeficiency disorder patients, E.Y. applied scales to patients with severe asthma. M.A. and M.Z.K applied scales to patients with malignancy. Y.A. applied scales to patients with hypertension and patients with cardiovascular system disease. M.S. applied scales to patients with diabetes mellitus. F.Ç^1^., G.A., and M.S. applied scales to healthcare workers. F.Ç^1^., O.K., F.Ç^5^. selected the references and extracted the data. F.Ç^1^, O.K., and ş.B. analyzed the data. F.Ç^5^., G.A., and M.S. contributed to the interpretation of the data. All authors contributed to the draft of the study and read and approved the final manuscript.

## List of abbreviations

ACEI: Angiotensin converting enzyme inhibitors
ARB: Angiotensin II receptor blocker
ARDS: Acute respiratory distress syndrome
AUC: Area under the curve
CI: Confidence interval
COVID-19: Coronavirus disease 2019
CID: Combined immunodeficiency
CVID: Common variable immune deficiency
CVS: Cardiovascular system
DM: Diabetes mellitus
DPP-4: Dipeptidyl peptidase-4
FIVE: Fear of illness and virus evaluation
HADS: Hospital anxiety and depression scale
HADS-A: Hospital anxiety and depression scale, anxiety subscale
HADS-D: Hospital anxiety and depression scale, depression subscale
HCC: Hepatocellular carcinoma
HT: Hypertension
NK: Natural killer
PID: Primary immunodeficiency disorder
SARS-CoV-2: Severe acute respiratory syndrome coronavirus-2
SCIG: Subcutaneous immunoglobulin

